# Awareness, utility and preferences of campus-based mental health services at tertiary institutions in Harare, Zimbabwe: a cross-sectional study

**DOI:** 10.1101/2025.08.08.25333276

**Authors:** Omega Mukubvu, Gamuchirai C Tonono, Ropafadzo L Mupinga, Chris Uriga, Yola A Machona, Shalom R Doyce, Edwin Mavindidze, Rufaro H. Mushonga, Beatrice K Shava, Sidney Muchemwa, Dixon Chibanda, Anotida R Hove, Jermaine Dambi

## Abstract

Mental health (MH) disorders are highly prevalent among university students, with multi-level impacts. Although campus-based mental health services are available, awareness and utilisation rates remain low. This study examined the awareness and utilisation of campus mental health services among Zimbabwean tertiary students, including the barriers, facilitators, and preferences that influence their access to support. This descriptive cross-sectional study recruited 1070 students from five tertiary institutions in Harare, Zimbabwe. Participants completed questionnaires evaluating awareness, utilisation, preferences, and barriers to accessing campus-based mental health services. Data were analysed using descriptive statistics and multivariate logistic regression. The average age of the participants was 21.7 (SD = 2.7). 76.5% of students were aware of campus-based MH services. Awareness of MH services was associated with familial history of MH conditions [AOR= 1.35 (95%CI: 1.05; 1.83), p=.05] and personal experience of a MH condition [AOR= 0.71 (95%CI: 0.52; 0.97), p=.030]. Only 16.5% of students had utilised campus-based MH services. High usage was associated with drug and substance use [AOR= 2.388 (95%: 1.227; 4.644), p= 0.01], availability of a psychologist [AOR = 1.69 (95%CI: 1.10; 2.59), p=.017] and availability of campus-wide MH workshops [AOR = 1.62 (95%: 1.00; 2.60), p=.049]. Key barriers to MH service utilisation included lack of resident/institutional MH service providers [AOR= 2.02 (95%CI: 1.16; 3.51), p=.013], past MH experiences [AOR= 1.53 (95%CI: 1.16; 2.02), p=.002], and friend’s history of MH condition [AOR= 1.44 (95%CI: 1.11; 1.86), p=.006]. Students preferred self-help services, individual therapy, and psychologists for MH support. Universities should promote MH awareness through effective awareness campaigns and workshops, provide tailored services that align with the students’ preferences, and ultimately create a supportive environment that fosters students’ overall mental health service utilisation and well-being.

## Introduction

Common mental disorders (CMDs), particularly depression and anxiety, are highly prevalent among university students. Approximately 35% of university students experience significant mental health (MH) problems in a lifetime [1,2]. The beginning of tertiary education signifies a transitional period for young adults, during which they face challenges such as making independent decisions about their personal lives and adapting to increased academic pressures [3]. The transitional period is characterised by an increased risk of CMDs, uncertainties about the future, potentially leading to a vicious cycle of poor MH, and decreased academic performance and social functioning [4–6]. Moreover, in low-resource environments, students who relocate from other places for tertiary education often encounter additional obstacles. These include limited social support and financial hardships, both of which elevate their risk of experiencing poor MH [7]. Therefore, early identification and treatment of CMDs are crucial for students to mitigate the adverse effects [8]. Failure to address CMDs in tertiary students can result in lower academic performance, dropouts, increased substance use, risky sexual behaviours, and social difficulties. These problems may persist into adulthood, resulting in various psychosocial and economic consequences [9–12]. Consequently, various campus-based mental health (CBMHS) interventions, including counselling services, digital interventions, referral pathways for stepped-up care, and health promotional initiatives, have been implemented to improve students’ mental well-being [13,14]. CBMHS serve as a crucial entry point for both preventive and curative care. These services (CBMHS) are potentially cost-effective and enhance accessibility to MH services for tertiary students [7,15]. However, several factors such as limited availability of specialised professionals, concerns about stigma, and privacy considerations can affect the uptake and utilisation of CBMHS [16–18]. For instance, a systematic review analysing 44 studies showed that the utilisation of MH services among universities ranged from as low as 13.7% to 68.7% [16]. However, in this systematic review, only one article was from Sub-Saharan Africa (SSA), with the majority of studies (n = 32) originating from the USA [16]. The lack of evidence from the SSA region is concerning, given the historically disproportionately huge MH care gap within the region [7,19].

Generally, the awareness and uptake of MH services in low- and middle-income countries is significantly lower compared to high-income countries [7,20,21]. A limited awareness and utilisation of the existing CBMHS exacerbate the high care gap within universities in SSA [7,22]. The key active ingredients for the optimal uptake or utilisation of CBMHS include increased awareness, positive attitudes, and perceptions towards the available MH services. The underutilisation is influenced by factors such as low mental health literacy (MHL), stigma, negative attitudes, peer labelling, lack of awareness, gender expectations, academic pressure, cultural beliefs, time constraints, confidentiality concerns, accessibility issues, low resources, self-reliance, and preference for support from family and friends [7,18,20,23–28]. For example, a Nigerian cross-sectional study (N=450) reported that 67% of students had never used MH services [29]. The low utilisation was attributed to poor health-seeking behaviours, limited social support, and insufficient information on available MH services [29]. Another cross-sectional study among South African tertiary students (N=28516) showed that 71.3% reported having an MH problem, yet only half (35.2%) had utilised on-campus MH services [21]. Adolescents and young adults are especially susceptible to CMDs but often hesitate to seek professional help due to various barriers to accessing MH services, such as stigma, negative attitudes towards treatment, beliefs, low resources and financial barriers [30]. Tertiary students seek MH services when assured of confidentiality, respect, and no judgment [7]. Factors such as positive past experiences, good MHL, encouragement from others, and belief in effective treatment also influence engagement with CBMHS [7,15]. Furthermore, the acceptance and utilisation of MH services by university students are closely linked to user attitudes and preferences regarding the mode of delivery of MH services [31,32]. For instance, studies have demonstrated disparities in the preferences for the mode of delivery of MH services. Some students prefer face-to-face over online mental health services, while others favour blended, i.e., a combination of both physical and online CBMHS [33,34]. Therefore, it is paramount to understand the MH service preferences of Zimbabwean students to inform the development of targeted MH services or modification of existing services. Importantly, Zimbabwe has also seen an exponential increase in the burden of CMDs in tertiary students, as in the general population. However, as with other low-resource settings, a huge mental care gap exists, and there have not been previous attempts to explore the uptake of the available CBMHS systematically in the Zimbabwean context. This study, thus, aimed to assess the utilisation and awareness of CBMHS among Zimbabwean tertiary students. We also sought to identify the barriers, facilitators, and preferences of tertiary students regarding the use of CBMHS. It is important to evaluate the utility of CBMHS, particularly to identify barriers, and understand students’ preferences to develop tailored interventions that address the mental health needs of tertiary students from low-resource settings, which are plagued with a disproportionately high MH care gap. Insights from this study could guide the design of bespoke interventions, including optimising the uptake of existing services to improve the mental well-being of university students.

## Materials and methods

### Ethics statement

Ethical approval was obtained from the Joint Parirenyatwa & the University of Zimbabwe, Faculty of Medicine and Health Sciences Research and Ethics Committee (JREC) JREC: 25/2025). Institutional approval was granted from the five tertiary institutions where the study was conducted. The study adhered to the principles outlined in the Declaration of Helsinki. For instance, written informed consent was obtained from each participant, and they were treated as autonomous agents, as they could withdraw from the study at any time without consequence.

### Study design and setting

This cross-sectional study was conducted from February 24, 2025, to April 28, 2025. Participants were recruited from five tertiary institutions, i.e. the University of Zimbabwe, the Women’s University in Africa, the Harare Institute of Technology, Harare Polytechnic, and the Catholic University of Zimbabwe. The study sites are all located in Harare, the capital city of Zimbabwe. Further, the five institutions have a combined enrolment of 34,557 students, approximately 30% of all tertiary students in Zimbabwe. At various institutions, different CBMHS are offered, such as campus-wide mental health awareness campaigns, psychotherapies, peer-led interventions, and referrals for stepped-up care.

### Participants

University students were recruited through a multistage sampling process. The first stage involved stratified proportionate sampling, where a proportional sample was selected from each of the five institutions. The stratum size was proportional to the university’s total intake/size relative to the study population. Thereafter, university students were conveniently recruited into the study. To be included, study participants had to be 18 years or older, enrolled as undergraduate students at the respective institutions, able to read and understand English, and voluntarily consent to participate in the study. Participants were excluded from the study if they were unable to give consent due to psychological conditions or intoxication.

### Sample size calculation

The sample size was based on estimates from a similar survey conducted to estimate the utilisation of on-campus MH services among Nigerian university students. The Nigerian study yielded a 36 % (p_0_=0.36) utilisation prevalence [35]. Assuming a comparable estimate of 31% in Zimbabwean students (p_1_=.31), a minimum of 1066 students were required at a 95% significance level (α =0.05) and 80% goal power (1-β). The sample size was calculated using Statistica Version 14.

### Data collection procedure

Prior to data collection, the researchers conducted a situational analysis to determine the range of mental health services available at all the institutions. Upon arrival, researchers approached university students in their lecture rooms and various public areas such as canteens and foyers. Researchers explained the aims and procedures to students. Participants were informed of their right to freely participate and withdraw from the study at any moment without any consequences. Volunteering students were identified, given consent forms to review, and given the opportunity to ask any questions about the study. They were then asked to sign once they had understood and agreed to participate. Once researchers obtained written informed consent, participants completed the questionnaire programmed on KoboCollect using electronic tablets. Study questionnaires were self-administered; however, researchers were nearby to offer help as needed.

### Data collection instruments

#### Participants characteristics questionnaire

Sociodemographic characteristics were elicited through an ad hoc questionnaire which captured participants’ age, sex, institution, faculty, year of study, religion, area of residence, alcohol intake, smoking status, financial adequacy and personal and familial history of MH conditions.

#### Awareness, utilisation and barriers questionnaires

Participants were asked to indicate their knowledge of the MH services available by selecting the services provided at their respective institutions. The utilisation of MH services was assessed via a questionnaire, which identified the 12-month prevalence of MH service utilisation and the likelihood of future use. MH services preferences were gathered by asking participants about their preferred location (on-campus or off-campus), delivery mode (in-person, online, self-help, or hybrid), and delivery agent (e.g. lay counsellor, psychiatrist, psychologist). Barriers to accessing MH services were measured using questions adapted from studies that aimed to determine the barriers to MH services in other populations. This questionnaire was primarily adapted from the Barriers to Mental Health Services Scale (BMHSS) [36]. The complete questionnaire was piloted among 35 participants to test its face validity, clarity, acceptability (respondent burden), and appropriateness. Changes were made to the questionnaire, including the omission of similar items, merging of questions, and rephrasing of questions to improve clarity and conciseness.

### Data analysis

Data were analysed using descriptive statistics to summarise the participants’ characteristics, MH utilisation, preferences, awareness and perceived barriers to utilisation of CBMHS. Bivariate and multivariate logistic regression were used, and adjusted odds ratios (AOR) and associated 95% confidence intervals were determined to identify factors associated with awareness, utilisation, preferences, and perceived barriers to utilisation of MH services among undergraduate students. All analyses were conducted at an α level of 0.05 using SPSS (Version 29).

## Results

### Participant characteristics

The study recruited 1,070 tertiary-level students from five institutions, with a mean age of 21.7 years (SD = 2.7). Most of the participants were: female (54.3%; n= 581), in their first year of study (43.0%; n = 460), studying engineering-related courses (31.9%; n= 341), full-time students (97.6%; n= 1044), Christian (91.0%; n = 974), resided with family (43.6%; n = 466), and rated their monthly finances as adequate for them (32.2%; n= 345). Also, most participants did not smoke (91.4%; n= 978), take alcohol (72.6%; n= 777), nor use any substances (95.4%; n= 1021). The lifetime prevalence of MH conditions was 35%, with a 12-month period prevalence of 66.7%. The most common conditions reported were depression (35.0%; n=300) and anxiety (21.8%; n=233). Lastly, most participants had a family member (46.1%; n=493) or a friend (49.0%; n=524), who had a history of a MH condition – See Table 1.

**Table 1:** Participant characteristics, N=1070.

| Variable | Attribute | Frequency, n (%) |
| --- | --- | --- |
| Gender | Female | 581 (54.3) |
|  | Male | 475 (44.4) |
|  | Other | 14 (1.3) |
| *Age | Mean (SD) | 21.7 (2.7) |
| Institution | Institution A | 149 (13.9) |
|  | Institution B | 106 (9.9) |
|  | Institution C | 286 (26.7) |
|  | Institution D | 120 (11.2) |
|  | Institution E | 409 (38.2) |
| Academic year | First | 460 (43.0) |
|  | Second | 261(24.4) |
|  | Third | 70 (6.5) |
|  | Fourth | 279 (26.1) |
| Faculty or course | Technical programs | 41 (3.9) |
|  | Arts And Humanities | 212 (19.8) |
|  | Health And Sciences. | 325 (30.3) |
|  | Commercials | 151 (14.1) |
|  | Engineering | 341(31.9) |
| Enrolment classification | Full-time student | 1044 (97.6) |
|  | Part-time student | 26 (2.4) |
| Religion | African Traditional Religion | 29 (2.7) |
|  | Christian | 974 (91.0) |
|  | Islam | 14 (1.3) |
|  | None/Atheist | 22 (2.1) |
|  | Other | 31(2.9) |
| Residence | On-campus accommodation | 212 (19.8) |
|  | Off-campus housing | 358 (33.5) |
|  | Stays with family | 466 (43.6) |
|  | Other | 34 (3.2) |
| Perceived financial adequacy | Very inadequate | 183 (17.1) |
|  | Inadequate | 241 (22.5) |
|  | Somewhat Inadequate | 245 (22.9) |
|  | Adequate | 345 (32.2) |
|  | Very adequate | 56 (5.2) |
| Alcohol Intake | No | 781781 (73.0) |
|  | Yes | 289289 (27.0) |
| Smoking status | No | 978 (91.4) |
|  | Yes | 92 (8.6) |
| Drug and substance use | No | 1021 (95.4) |
|  | Yes | 49 (4.6) |
| Ever experienced a mental health condition | No | 695 (65.0) |
|  | Yes | 375 (35.0) |
| Experienced a mental health condition in the past twelve months | No | 121 (32.3) |
|  | Yes | 254 (66.7) |
| Type of mental health condition experienced in the last 12 months | Depression | 300 (28.0) |
|  | Anxiety | 233 (21.8) |
|  | Post-traumatic stress disorder | 99(9.3) |
|  | Bipolar disorder | 34(3.2) |
|  | Substance and drug abuse | 20(1.9) |
|  | Other | 19(1.8) |
| Family history of mental health condition | No | 577(53.9) |
|  | Yes | 493(46.1) |
| Having a friend with a mental health condition | No | 546(51.0) |
|  | Yes | 524(49.0) |

### Awareness of campus-based mental health services

Over three-quarters of the participants (76.5%; n = 819) reported being aware of any CBMHS. Of those aware of the existing on-campus MH services, most students were aware of the existence of face-to-face counselling services (56.5%; n = 463) and peer counselling services (19.8%; n = 212). The least recognised MH services were online services/telehealth, with only 9.5% (n = 102) and 7.4% (n = 79) aware of external MH service providers (see Table 2).

**Table 2:** Awareness of on-campus mental health services.

| On-campus mental health services available per institution | Attribute | Frequency, n (%) |
| --- | --- | --- |
| Online service/ telehealth | No | 968(90.5) |
|  | Yes | 102(9.5) |
| Face-to-face counselling services | No | 607(56.7) |
|  | Yes | 463(43.3) |
| A campus-based psychologist | No | 878(82.1) |
|  | Yes | 192(17.9) |
| Campus-based chaplain | No | 889(83.1) |
|  | Yes | 181(16.9) |
| The university has partnered with an off-campus mental health service provider. | No | 991(92.6) |
|  | Yes | 79(7.4) |
| Mental health workshops/talks | No | 918(85.8) |
|  | Yes | 152(14.2) |
| Mental health support groups | No | 940(87.9) |
|  | Yes | 130(12.1) |
| Peer educators | No | 858(80.2) |
|  | Yes | 212(19.8) |
| Mental health awareness campaign programs | No | 875(81.8) |
|  | Yes | 195(18.2) |
| None | No | 819(76.5) |
|  | Yes | 251(23.5) |

**Table 3:**
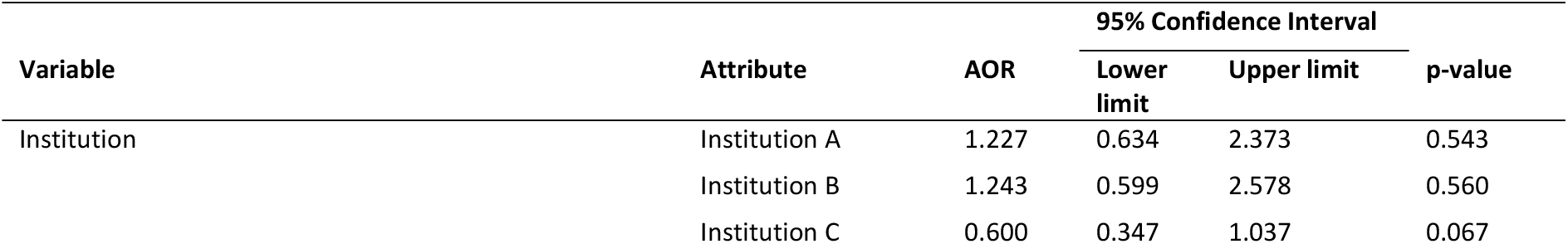
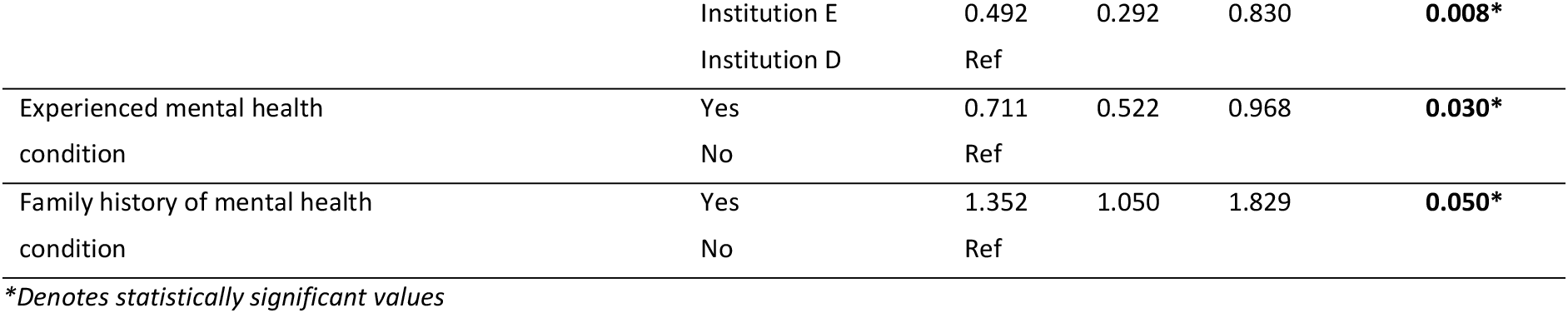
Factors associated with awareness of campus-based mental health services.

| Variable | Attribute | AOR | 95% Confidence Interval |  | p-value |
| --- | --- | --- | --- | --- | --- |
|  |  |  | Lower limit | Upper limit |  |
| Institution | Institution A | 1.227 | 0.634 | 2.373 | 0.543 |
|  | Institution B | 1.243 | 0.599 | 2.578 | 0.560 |
|  | Institution C | 0.600 | 0.347 | 1.037 | 0.067 |
|  | Institution E | 0.492 | 0.292 | 0.830 | **0.008\*** |
|  | Institution D | Ref |  |  |  |
| Experienced mental health condition | Yes | 0.711 | 0.522 | 0.968 | **0.030\*** |
|  | No | Ref |  |  |  |
| Family history of mental health condition | Yes | 1.352 | 1.050 | 1.829 | **0.050\*** |
|  | No | Ref |  |  |  |

\*Denotes statistically significant values

The S1 Table presents the crude odds ratios of factors associated with awareness of on-campus MH services, while **Error! Reference source not found**. displays the adjusted analysis. After adjustment for confounding and covariance, only institution, experience and familial history of mental conditions were significantly associated with awareness of CBMHS. Compared to students from D, students at institution E were 50.8 % less likely to be aware of the available MH services (AOR= 0.492 (95%:0.292; 0.830), p=.008). Also, participants who experienced a MH condition were 28.9% less likely to be aware of available MH services than those who had not experienced a MH disorder (AOR= 0.711 (95%:0.522; 0.968), p=.030). Last, participants with a familial history of MH conditions were almost 1.4 times more likely to be aware of available MH services than those without a familial history of MH conditions (AOR= 1.352 (95%: 1.050; 1.829), p=.05).

### On-campus mental health service utilisation

Only a few students (16.5%; n = 177) had utilised CBMHS since they were enrolled at their institutions; of these, more than half (55.4%; n = 98) reported utilisation in the past 12 months. Out of the students who had utilised MH services in the past 12 months, most (6.7%; n = 72) reported that they had rarely used them. Most respondents were likely to recommend the on-campus MH services to others (10.8%; n = 116). Lastly, most students felt they were likely to utilise MH services in the future (54.0%; n= 578). See Table 4.

**Table 4:** Frequencies of utilisation of on-campus mental health services, N=1070.

| Variable | Attribute | Frequency, n (%) |
| --- | --- | --- |
| Utilisation since enrolment | No | 893 (83.5) |
|  | Yes | 177 (16.5) |
| Utilisation of on-campus mental health services in the past 12 months | No | 79 (44.6) |
|  | Yes | 98 (55.4) |
| Frequency of utilisation of on-campus mental health services in the last year | Only once | 34 (3.2) |
|  | Rarely (yearly) | 72 (6.7) |
|  | Occasionally (month) | 58 (5.4) |
|  | Regularly(week) | 13 (1.2) |
| Likelihood of recommendation of on-campus mental services to others | Very unlikely | 25 (2.3) |
|  | Unlikely | 14 (1.3) |
|  | Indifferent | 22 (2.1) |
|  | Likely | 58 (5.4) |
|  | Very likely | 58 (5.4) |
| Likelihood of future usage of on-campus mental services | Very unlikely | 249 (23.3) |
|  | Unlikely | 243 (22.7) |
|  | Somewhat likely | 226 (21.1) |
|  | Likely | 241(22.5) |
|  | Very likely | 111(10.4) |

The **S2 Table** presents the crude odds ratios of factors associated with utilising on-campus MH services, while Table 5 displays the adjusted analysis. After adjusting for other variables, institution, availability of MH services, preferred MH professional, and drug and substance use remained significantly associated with MH service utilisation. Students from institutions A, B, C and E were less likely to utilise CBMHS compared to students from institution D, with reductions in odds of 79.4% [AOR=.206 (95% CI: 0.125; 0.337), p<.001], 82.4% [AOR=.176 (95%CI: 0.121; 0.256), p<.001], 81.3% [AOR=.187 (95%CI: 0.131; 0.268), p<.001], and 81.1% [AOR=.199 (95%CI: 0.114; 0.348), p<.001], respectively. Further, students with a history of drug and substance use were 2.4 times more likely to utilise MH services compared to those without [AOR =2.388 (95%CI: 1.227; 4.644), p< 0.001]. Next, students with access to support groups, MH service providers, psychologists, or workshops on their campus were significantly more likely to utilise MH services compared to those without, with increased odds of 2.4 times [AOR= 2.069 (95 %CI: 1.26; 3.395), p=.004],1.8 times [AOR= 1.838 (95%CI: 1.009;3.348), p=.047], 1.7 times [AOR= 1.686 (95%CI:1.098; 2.59), p=.017] and 1.7 times [AOR= 1.615 (95%CI: 1.003; 2.6), p=.049] respectively. Last, participants who preferred psychologists or psychiatrists as their MH service providers were significantly less likely to utilise MH services, with reduction in odds of 47.4% [AOR =0.526 (95%CI: 0.384; 0.72), p <.001] and 41.2% [AOR =0.588: (95%CI: 0.358; 0.966), p= 0.036], respectively. See Table 5

**Table 5:** Factors associated with the utilisation of campus-based mental health services.

| Variable | Attribute | Adjusted<br>Odds ratio | 95% Confidence interval |  | p-value |
| --- | --- | --- | --- | --- | --- |
|  |  |  | Lower limit | Upper limit |  |
| Institution | Institution A | 0.206 | 0.125 | 0.337 | <.001* |
|  | Institution B | 0.199 | 0.114 | 0.348 | <.001* |
|  | Institution C | 0.176 | 0.121 | 0.256 | <.001* |
|  | Institution E | 0.187 | 0.131 | 0.268 | <.001* |
|  | Institution D | Ref |  |  |  |
| Drug and substance use | Yes | 2.388 | 1.227 | 4.644 | < 0.001* |
|  | No | Ref |  |  |  |
| Available mental health services: |  |  |  |  |  |
| Psychologist | Yes | 1.686 | 1.098 | 2.59 | 0.017* |
|  | No | Ref |  |  |  |
| External mental health service provider | Yes | 1.838 | 1.009 | 3.348 | 0.047* |
|  | No | Ref |  |  |  |
| Workshops | Yes | 1.615 | 1.003 | 2.6 | 0.049* |
|  | No | Ref |  |  |  |
| Support groups | Yes | 2.069 | 1.26 | 3.395 | 0.004* |
|  | No | Ref |  |  |  |
| Preferred professional: |  |  |  |  |  |
| Psychologist | Yes | 0.526 | 0.384 | 0.72 | <.001* |
|  | No | Ref |  |  |  |
| Psychiatrist | Yes | 0.588 | 0.358 | 0.966 | 0.036* |
|  | No | Ref |  |  |  |

\*Denotes statistically significant values

### Perceived barriers to utilisation

The most endorsed barriers to accessing CBMHS included: fear of being seen as weak for having a MH condition (46.7%, n=499), feelings of shame or embarrassment (44.2%, n=472), past bad experiences with CBMHS (67.9, n=727), the belief that professional help would likely be ineffective (61.3%, n=656), being too unwell to ask for help (56%, n=599), and concerns about cost (42.7%, n=457) – See Table 6.

**Table 6:** Barriers to access to on-campus mental health services, N=1070.

| Domain | Item | Strongly disagree, n (%) | Disagree, n (%) | Agree, n (%) | Strongly agree, n (%) |
| --- | --- | --- | --- | --- | --- |
| Stigma-related barriers | Concern about what my family might think, say, do or feel | 146(13.6) | 190(17.8) | 590(55.1) | 144 (13.5) |
|  | Concern that I might be seen as weak for having a mental health problem | 175(16.4) | 324(30.3) | 390(36.4) | 181(16.9) |
|  | Feeling embarrassed or ashamed | 159(14.9) | 313(29.3) | 414(38.7) | 184(17.2) |
|  | Concern about what my friends might think, say or do | 132(12.3) | 294(27.5) | 456(42.6) | 188(17.6) |
|  | Concern about what students might think, say or do | 164(15.3) | 269(25.1) | 454(42.4) | 183(17.1) |
|  | I am worried about my privacy and confidentiality if I use campus-based mental health services | 113(10.6) | 210(19.6) | 435(40.7) | 312(29.2) |
| Attitudinal-related barriers | Thinking the problem would get better by itself | 152(14.2) | 325(30.4) | 436(40.7) | 157(14.7) |
|  | Wanting to solve the problem on my own | 111(10.4) | 313(29.3) | 450(42.1) | 196(18.3) |
|  | Preferring to get help from family or friends | 128(12.0) | 287(26.8) | 531(49.6) | 124(11.6) |
|  | Dislike of talking about my feelings, emotions or thoughts | 110(10.3) | 270(25.2) | 463(43.3) | 227(21.2) |
|  | Thinking that professional care probably would not help | 178(16.6) | 478(44.7) | 308(28.8) | 106(9.9) |
|  | Having had previous bad experiences with campus-based mental health services | 258(24.1) | 469(43.8) | 243(22.7) | 100(9.3) |
| Instrumental-related barriers | Not being able to afford the financial costs involved | 98(9.2) | 249(23.3) | 524(49.0) | 199(18.6) |
|  | Having no one who could help me get professional care | 124(11.6) | 333(31.1) | 476(44.5) | 137(12.8) |
|  | Being unsure where to go to get professional care | 90(8.4) | 291(27.2) | 539(50.4) | 150(14.0) |
|  | Failing to find time to visit campus-based mental health services because of academic commitments | 105(9.8) | 294(27.5) | 502(46.9) | 169(15.8) |
|  | Being too unwell to ask for help | 170(15.9) | 429(40.1) | 377(35.2) | 94(8.8) |

The S3 Table presents the crude odds ratios of factors associated with barriers to accessing on-campus MH services, while Table 7 displays the adjusted analysis. After multivariate adjustments, only institution, experience with MH condition(s), availability of certain on-campus MH services, and lifetime utilisation of on-campus MH services were associated with barriers to service utilisation. First, students at institutions with an MH service provider were twice as likely to perceive barriers to utilising CBMHS compared to students at an institution without an MH service provider (AOR = 2.016 [95% CI: 1.157; 3.514], p =.013). Second, students who had participated in MH service workshops were 1.6 times more likely to perceive barriers to utilisation of MH services compared to those who did not participate in CBMHS workshops [AOR=1.615 (95%: 1.089; 2.395), p =.017]. Third, participants who had experience of a MH condition in their lifetime were 1.5 times more likely to perceive barriers to CBMHS than students who had never experienced a MH condition in their lifetime [AOR = 1.533 (95%:1.164; 2.019), p =.002]. Fourth, participants with a friend diagnosed with a MH condition were 1.4 times more likely to face barriers to utilisation of CBMHS than those who had no friend diagnosed with a MH condition [AOR = 1.437 (95%: 1.109; 1.863), p =.006]. Fifth, compared to students at D, students at institution C are 53.6% more protected from experiencing the barriers to accessing MH services [AOR=0.464 (95%:0.295; 0.728), p<.001] while students at B are 62% more protected from experiencing barriers to accessing MH services [AOR=0.380 (95%:0.219; 0.661), p<.001] and students at institution A are 43.5% more protected from experiencing barriers to accessing MH services [AOR= 0.565 (95%: 0.335; 0.954), p=.033]. Lastly, students who had utilised CBMHS since enrolment were 40.9% less likely to perceive barriers to utilisation of CBMHS compared to those who had never utilised CBMHS, [AOR = 0.591 (95%: 0.418; 0.835), p = 0.008].

**Table 7:**
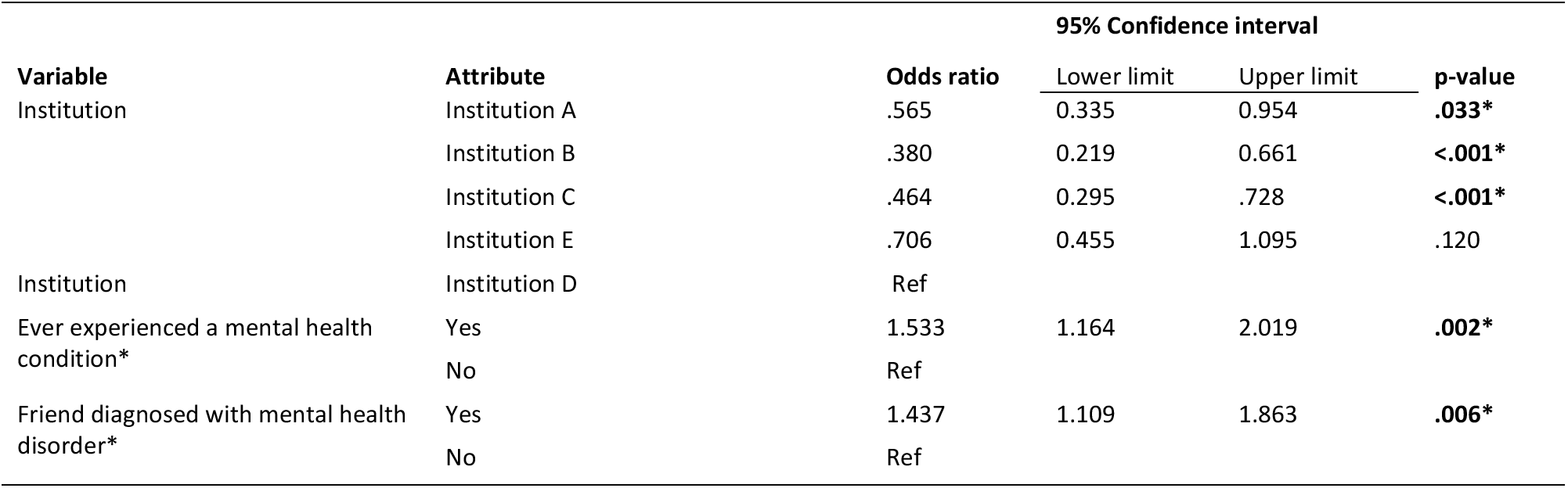
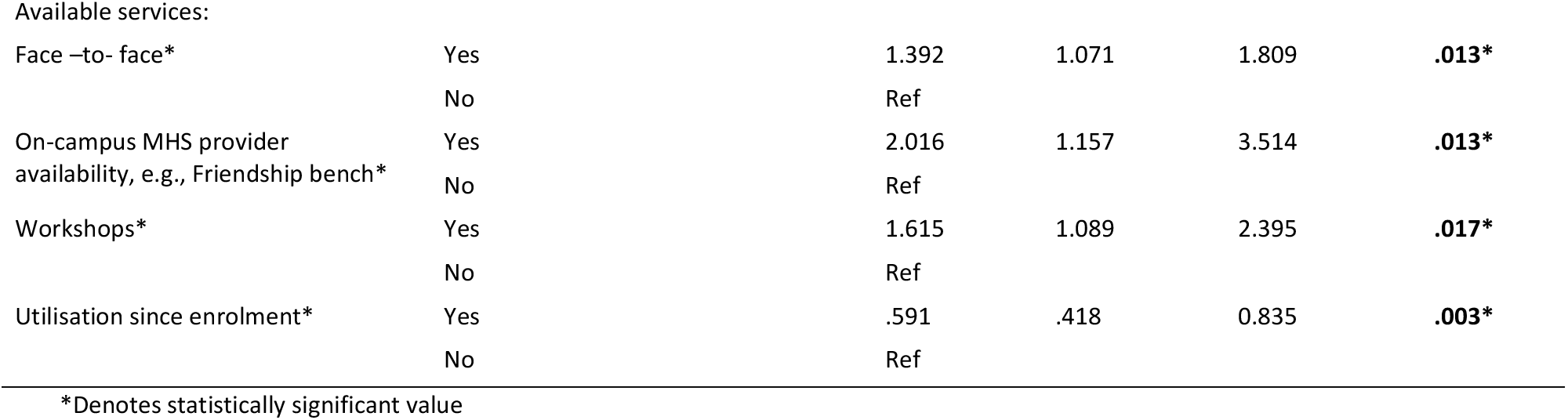
Factors associated with barriers to the utilisation of campus-based mental health services.

**Preferences**

Most students (52.6%; n = 563) indicated a preference for individual on-campus therapy. Two-thirds of the students preferred in-person, campus-based services (65.4%; n = 603). Lastly, more than three-quarters of the students (82.7%; n = 885) chose self-help as a viable source of MH support during a crisis, and psychologists were the leading preferred MH service providers (55.2%, n=591) - See Table 8.

**Table 8:** Students’ preferences for mental health services.

| Preference Category | Variable | Attribute | Frequency, n (%) |
| --- | --- | --- | --- |
| Location and intervention delivery format | Delivery mode | Individual therapy on campus | 563 (52.6) |
|  |  | Individual therapy off-campus | 356 (33.3) |
|  |  | Group therapy on campus | 272 (25.4) |
|  | Location and mode | In-person campus-based | 603 (65.4) |
|  |  | In-person off-campus | 483 (45.1) |
|  |  | Digital | 440 (41.1) |
| Preferred mental health professional | Psychologist | No | 479 (44.8) |
|  |  | Yes | 591 (55.2) |
|  | Psychiatrist | No | 895 (83.6) |
|  |  | Yes | 175 (16.4) |
|  | Counsellor | No | 583 (54.5) |
|  |  | Yes | 487 (45.5) |
|  | Peer counsellor | No | 757 (70.7) |
|  |  | Yes | 313 (29.3) |
|  | Dean of Students | No | 970 (90.7) |
|  |  | Yes | 100 (9.3) |
|  | Janitor | No | 1019 (95.2) |
|  |  | Yes | 51(4.8) |
|  | Social worker | No | 538 (50.3) |
|  |  | Yes | 532 (49.7) |
|  | Self-help service | No | 185 (17.3) |
|  |  | Yes | 885 (82.7) |

## Discussion

This study examined the awareness and utilisation of CBMH among Zimbabwean tertiary students, including the barriers, facilitators, and preferences that influence their access to support. 76.5% of the participants were aware of any CBMHS, yet only 16.5% of students utilised these. Awareness and use were linked to personal or family MH condition history, past experiences with on-campus services, support availability, and institutional factors.

### Awareness of campus-based mental health services and associated factors

In this study, results are consistent with previous research, indicating that a significant proportion of students are aware of available MH resources [37,38]. A scoping review yielded awareness rates in the 13-54% range [39]. Similarly, a cross-sectional study conducted in the United States (N=292) revealed that two-thirds of the tertiary students were aware of the MH services available on campus [40]. In the United States study, although students were informed about existing services during first-year orientation, the campaign messages lacked crucial details about the type and location of services, instead providing only general information about their existence [40]. In this study, discrepancies were found in awareness of CBMHS among the five universities recruited. This difference could stem from variable institutional policies and infrastructure, including health promotional efforts. For instance, universities with higher levels of engagement in promoting MH services often use targeted campaigns, workshops, or partnerships with MH organisations, which can significantly boost awareness levels [38]. Also, the accessibility and visibility of these services on campus could influence awareness levels. For example, clear signage, centralised help centres, or integration of MH resources into student wellness services might make services better known across campuses [41–43]. Cultural stigma around MH can prevent students at some institutions from seeking or discussing support services [43]. Therefore, universities can enhance student support by implementing campus MH policies, improving resource accessibility, fostering partnerships, raising awareness, reducing stigma, and regularly monitoring and revising the policies to ensure their effectiveness and support student well-being and academic success [44]. Also, in this study, participants with a familial history of MH disorders were more likely to be aware of available mental health services. These results are consistent with an Egyptian cross-sectional study (N=1740) among undergraduates, which found that participants with a familial history of MH issues were 1.8 times more likely to be aware of MH services than those without a familial history of MH conditions [45]. Exposure to mental health conditions in families increases awareness of services and treatment options through open discussions and direct experiences [46]. MH awareness and knowledge are closely linked to personal experience [46], aligning with social learning theory, which asserts that observation influences behaviour [47].

Furthermore, participants who experienced a MH condition were 28.9% less likely to be aware of available CBMHS than those who had not experienced a MH condition. This suggests that individuals who have experienced a MH condition may be receiving services elsewhere (e.g. public hospitals, private mental health care), resulting in them not looking for or considering campus-based services. Alternatively, students with experience of MH condition may be focusing on managing their condition rather than actively seeking information about available services. They might perceive stress or anxiety related to academics as a normal aspect of college life, resulting in the normalisation of distress among students, and this has been observed in other studies [43]. However, these results diverge from past studies, for example, an American cross-sectional study (N=266) found that students who had experienced MH conditions were more likely to be knowledgeable of CBMHS (*B* =.145, *t* = 2.440, *p* =.015) [48]. The US study suggests that personal experience enhances the knowledge of existing support services [48]. The experiential learning theory suggests that awareness of CBMHS can be shaped by direct experience and reflections on MH services and campaigns, ultimately influencing their knowledge and understanding of MH care [49]. Elsewhere, Bangladeshi university students (N=2036) with good MH were less likely to have higher awareness of MH services than those with poor MH (OR 0.72, 95%CI:.60–.87, p<.01) [50]. Further studies are needed to understand the link between awareness of existing on-campus MH services and the experience of a MH condition. Nevertheless, the variable level of awareness of existing on-campus MH services across the study sites underscores the critical need for universities to prioritise continuous, universal/campus-wide health promotional initiatives, including increasing coverage of MH services to increase awareness, as this is a critical step towards reducing the burden of disease through preventative and health promotive approaches [45,51].

### Utilisation of campus-based mental health services and associated factors

Our study reflects low utilisation of CBMHS. Similarly, a systematic review and meta-analysis of undergraduate students from 10 European studies showed a utilisation prevalence range from 13.1% to 68.1% [52]; our study aligns with the lower range. Similarly, an Ethiopian cross-sectional study of 3,240 undergraduate students found that only 28.7% utilised MH services, despite a high prevalence of mental distress in this setting [46]. Low utilisation in the Ethiopian study was attributed to a lack of awareness and perceived barriers to utilisation [46]. Similarly, a Nigerian study of 450 undergraduate students reported a lifetime prevalence of MH service use of 36%, with cost concerns and stigma associated with seeking help driving he low utilisation [35]. In the Nigerian study, about 60% of participants rarely used MH services, and 46% were unlikely to use them in the future [35]. Suboptimal uptake and reluctance to use existing services are areas of concern. These issues may stem from low awareness, preference mismatch, previous dissatisfaction, personal characteristics, and fear of stigma. Consequently, individual students may turn to alternative support systems, such as peers, family, and self-management [43,53,54].

Low utilisation is multifaceted and is driven by various factors. In this study, discrepancies were found in the utilisation of CBMHS among the recruited universities. Some students at the recruited universities were more aware of MH services, increasing their likelihood of seeking help. Secondly, variations in institutional culture, policies, or student characteristics may influence MH service use [51,55]. For example, participants who preferred psychologists or psychiatrists as their service providers were 47.4% and 41.2% respectively less likely to utilise CBMHS. Students may avoid seeking professional help for mental health concerns due to believing their issues aren’t serious enough, preferring self-management, getting help elsewhere, lacking information about available resources, or not having access to their preferred services on campus [56]. Also, the shortage of these specialist services within our context may explain the observed association, as the unavailability and inaccessibility are associated with long waiting times and resultant lower utilisation. However, in a Nigerian cross-sectional study (N=450), participants who preferred psychiatrists as their service providers were more likely to utilise CBMHS [35]. Psychiatrists were viewed as specialised experts providing a comprehensive assessment and treatment of mental disorders, which can instil confidence in students that they can receive thorough care, hence increasing their likelihood to seek help [35]. Our study also observed that students with a history of drug and substance abuse were 2.4 times more likely to utilise campus-based MH services. This suggests that prior experiences with substance use may lead to a greater need for MH support, given the multiple impacts of substance use on MH [57]. Similarly, a cross survey of South African university students (N=1402) found that students with a history of drug and substance use were almost three times more likely to utilise MH services compared to those without [AOR= 2.89 (95%: 1.11; 7.50) p<.05] [58].

### Perceived barriers to the utilisation of campus-based mental health services and associated factors

In this study, students experienced several barriers to accessing CBMHS, with stigma-related barriers, particularly the perspectives of family or friends concerning MH and distress disclosure. Most of the participants (67.9%) cited previous bad experiences with CBMHS as an intrusive barrier. Negative experiences can undermine trust, diminishing confidence in the services provided. Additionally, students may be reluctant to seek help again due to the fear that they might encounter a similar negative experience in the future. [59]. Notably, 61.3% of participants also expressed reluctance to seek professional help, believing that professional care would not help. This may be attributed to a lack of awareness of MH care and treatment options [60]. Additionally, students may feel uncomfortable sharing their distress with professionals as compared to family and friends [35,54]. Furthermore, being too unwell to ask for help was a significant barrier, with almost half of the participants highlighting this issue. This may be because students suffering from CMDs may experience physical symptoms, such as fatigue, low energy, and irritability, which have a broad impact on their quality of life and motivation to seek help [62, 63].

In this study, students who have a family member or friend with a MH condition are more likely to perceive barriers to using CBMHS, often due to negative experiences shared by their loved ones. While this familiarity can heighten awareness and potentially help overcome barriers such as stigma and lack of knowledge, it may also lead to increased reluctance to seek help, stemming from fears of facing similar challenges [35,61,62]. Therefore, it is crucial to provide targeted education and students with second-hand lived experience of MH condition(s) in MH treatment efforts to reduce stigma and attitude-related barriers. Also, students who attended mental health workshops were more likely to face barriers to the utilisation of CBMHS. This finding is unexpected because workshops typically aim to inform students about available mental health services, including where and how to access them, reduce stigma associated with seeking help, and provide a supportive environment that enhances uptake of existing services [63]. Conversely, workshops provide students with a comprehensive understanding of the challenges encountered by individuals who use MH services, which may inadvertently deter them from seeking care [64]. Additionally, ineffective workshop delivery and consequent dissatisfaction with the provided information may lead to a decline in the utilisation of existing campus services [64]. Nevertheless, further studies, including qualitative inquiry, are required to understand better the relationship between workshop attendance and the barriers to utilising campus-based mental health services.

We also observed that students who utilised CBMHS since enrolment were less likely to face barriers. Utilisation of services can increase familiarity with available CBMHS and what they entail, enhancing service uptake [59]. Engaging with mental health services early allows students to recognise and manage issues before they escalate, which may result in better outcomes and fewer obstacles to future use of available support services [45,55,56]. Significantly, the accessibility and convenience of CBMHS can minimise perceived access barriers, particularly compared to off-campus alternatives, which may encourage future use [65]. Also, students at institutions with resident partner MH service providers, such as Friendship Bench, perceived more barriers to utilisation than those without. This was unexpected, as other studies have shown that onsite external partners facilitate early access to services, including destigmatising mental healthcare access [55,66]. However, students may find it challenging to navigate the system, particularly if they are hesitant to disclose their distress or lack awareness of available support, which can result in lower utilisation of services [43]. Also, a limited understanding of the partnership’s role, benefits or services might contribute to perceived barriers, and students may also have different expectations about what MH services should entail [35,51]. Additionally, our study suggests that a lack of awareness about the services and their benefits might contribute to this perception, as 61.3% of participants believed professional help would not be effective. This finding highlights the importance of not only providing MH services but also ensuring they meet students’ needs and expectations. Tertiary institutions should proactively address barriers to MH service utilisation by implementing awareness campaigns that educate students on early identification of mental health symptoms and available campus-based services [38]. Additionally, institutions must invest in service evaluations to ensure their offerings meet students’ needs, ultimately fostering a supportive environment and encouraging students to seek help when necessary [43,63].

### Preferences of campus-based mental health services

In our study, participants showed a greater acceptance of in-person therapy conducted on campus, followed by digital therapy, with blended therapy (a combination of physical and online sessions) being the least preferred. This finding aligns with the results from a multi-centre cross-sectional study of German university students (N = 1,376), which revealed a preference for in-person MH services. Specifically, in the German study 76% of the students preferred off-campus and 71% preferred on-campus services, as opposed to digital therapy (19%) and blended therapy (9%) [67]. Similarly, a Mexican cross-sectional study of first-year students (N = 7849) found that 38% of students preferred in-person therapy over other services [68]. Students may prefer in-person on-campus services because they offer a convenient, accessible, and supportive environment that fosters face-to-face interaction, greater therapeutic alliance, immediate support and a sense of community and security [69]. Also, most students preferred individual therapy over group therapy (85.9% vs. 59.7%). These results are comparable to those of a German cross-sectional cohort study (N = 1,376), which found that 76% preferred individual therapy and 67% opted for group therapy [67]. The German study found that preferences for individual therapy were due to embarrassment, stigma, and beliefs about effectiveness: reasons echoed in our study. Most participants (82.7%) in the study showed a strong preference for self-help services, despite their absence across five tertiary institutions. This highlights a growing inclination towards autonomous MH care [54]. Self-help services, lauded for their convenience and alignment with high literacy rates among tertiary students, present an accessible opportunity to bridge the care gap, especially in an increasingly digitalised world [54]. Also, more students opted for peer counsellors as their MH service provider of choice over professionals (e.g. psychiatrists). This is comparable to a US study (N = 14,175) in which counselling services were preferred from non-professionals, such as friends (70.5%) [41]. The preference for peer counsellors might be attributed to their relatability, approachability, and shared experiences, which can offer comfort and understanding in a non-clinical setting [70]. Given the recommendation by the WHO to task-shift care, tertiary institutions should prioritise equipping peer counsellors with the necessary knowledge to help students increase utilisation [70].

### Study strengths and limitations

To our knowledge, this study is one of the few that have examined the awareness, perceived utility, and barriers to accessing on-campus mental health services in Sub-Saharan Africa. Our study offers valuable insights into improving the utilisation and reach of mental health services in tertiary institutions. Also, recruiting a large sample across five tertiary institutions increases the study’s internal and external validity. However, the application of convenience sampling could have resulted in recruitment or selection bias. Ideally, participants would have been randomly selected; however, this was not feasible due to resource limitations. The collection tools employed were not formally validated within our context, which may have led to measurement and misclassification biases. Future studies should address these limitations by validating the tools formally to ensure reliability and accuracy before conducting research.

## Conclusion

Our study revealed a high level of awareness and low utilisation of campus-based mental health services among tertiary-level students. The main barriers to using these services were related to attitudes and a lack of information. Tertiary institutions should promote mental health awareness through tailored, campus-wide health initiatives that direct students to the available mental health services. Campus-based mental health services must be aligned with student preferences, and there should be ongoing assessments of their acceptability to foster a supportive environment for students’ well-being. Lastly, there is a need to leverage digital technologies to offer self-help mental health services and bridge the care gap within universities.

## Data Availability

The dataset has been deidentified and submitted as a supporting file in excel format.

## Acknowledgements

The researchers would like to thank the participants who took part in the study for their time and contributions. We would also like to extend our sincere gratitude to the institutions where the research was conducted for the support provided.

